# Cortico-Cerebellar Neurodynamics during Social Interaction in Autism Spectrum Disorder

**DOI:** 10.1101/2022.10.17.22281189

**Authors:** Fleur Gaudfernau, Aline Lefebvre, Denis-Alexander Engemann, Amandine Pedoux, Anna Bánki, Florence Baillin, Benjamin Landman, Frederique Amsellem, Thomas Bourgeron, Richard Delorme, Guillaume Dumas

**Affiliations:** Human Genetics and Cognitive Functions, Institut Pasteur, UMR 3571 CNRS, University Paris Diderot, Paris, France; Inria, HeKA, PariSantéCampus, Paris, France; Inserm, Centre de Recherche des Cordeliers, Sorbonne Université, Université de Paris Cité, Paris, France; Department of Child and Adolescent Psychiatry, Robert Debré Hospital, APHP, Paris University, Paris, France; Roche Pharma Research and Early Development, Neuroscience and Rare Diseases, Roche Innovation Center Basel, F. Hoffmann–La Roche Ltd., Basel, Switzerland; Université Paris-Saclay, Inria, CEA, Palaiseau, France; Research Unit Developmental Psychology, Department of Developmental and Educational Psychology, Faculty of Psychology, University of Vienna, Vienna, Austria; Université de Montréal, Department of Psychiatry, Montréal, QC, Canada; CHU Sainte-Justine Centre de Recherche, Precision Psychiatry and Social Physiology laboratory, Montreal, QC, Canada

**Keywords:** Neurodevelopment, Social Cognition, Theta Oscillations, Cerebellum, Digital Psychiatry, Autism

## Abstract

**Background:** Exploring neural network dynamics during social interaction could help to identify biomarkers of Autism Spectrum Disorders (ASD). Recently, the cerebellum, a brain structure that plays a key role in social cognition, has attracted growing interest. Here, we investigated the electrophysiological activity of the cortico-cerebrum network during real-time social interaction in ASD. We focused our analysis on theta oscillations (3-8 Hz), which have been associated with large-scale coordination of distant brain areas and might contribute to interoception, motor control, and social event anticipation, all skills known to be altered in ASD.

**Methods:** We combined the Human Dynamic Clamp, a paradigm for studying realistic social interactions using a virtual avatar, with high-density electroencephalography (HD-EEG). Using source reconstruction, we investigated power in the cortex and the cerebellum, along with coherence between the cerebellum and three cortical areas, and compared our findings in a sample of participants with ASD and with typical development (TD) (n = 140). We developed an open-source pipeline to analyse neural dynamics at the source level from HD-EEG data.

**Results:** Individuals with ASD showed a significant increase in theta band power during social interaction compared to resting state, unlike individuals with TD. In particular, we observed a higher theta power over the cerebellum and the frontal and temporal cortices in the ASD group compared to the TD group, alongside bilateral connectivity alterations between the cerebellum and the sensorimotor and parietal cortices.

**Conclusions:** This study uncovered ASD-specific alterations in the theta dynamics, especially in a network between the cerebellum and social-associated cortical networks.

## INTRODUCTION

Autism Spectrum Disorders (ASD) are neurodevelopmental disorders characterized by impaired social interactions, and repetitive, stereotyped behaviors (1). As diagnosis of ASD primarily relies on clinical assessment, identification of robust biomarkers, specifically on neural dynamics during social interaction, could help achieve earlier and more consistent diagnosis (2). Magnetoencephalography (MEG) and electroencephalography (EEG) are optimal for functional analysis (3,4), thanks to their millisecond time resolution and ability to explore large frequency bands - such as 0.5-60 Hz for EEG *vs* 0.01-0.1 Hz in functional magnetic resonance imaging (fMRI). The majority of quantitative EEG studies in ASD focused on brain activity during resting state, suggesting oscillatory anomalies that draw a U-shaped profile, with greater relative power in low frequency and high frequency bands (5). Although some studies have started investigating brain signals during action observation (6,7), no EEG study has been conducted during real-time social interaction in ASD (8).

We recently developed a novel paradigm, the Human Dynamic Clamp (HDC) for studying real-time social interactions based on interpersonal coordination between a human and a Virtual Partner (VP) (9). People coordinated hand movements with the observed movements of a virtual hand, the parameters of which depended on the real-time input from the participant’s own movements. Neural dynamics could be recorded in parallel with high-density (HD)-EEG, thus providing insights into the dynamics of real-time social interactions in a controlled experimental setting, since the social interaction variability was only dependent on the participant (10). The HDC task in participants with typical development (TD) revealed local recruitment in the theta band activity in the motor areas during movement execution and in the right parietal areas during coordination with the virtual hand, as well as distributed connectivity across the antero-posterior network during intention attribution to the VP (11). Recent empirical work with the HDC in ASD provided preliminary evidence of a relationship between sensory-motor and socio-cognitive impairments (12).

Recently, the link between ASD and the cerebellum, which has a critical role in social interactions (13), has attracted growing interest. The cerebellum contributes to the understanding and imitating of others’ actions by observing their body (14), and mentalizing, *i*.*e*. inferring another person’s state of mind (15). It is also involved in the neural encoding of rhythmic processes (16). While cerebellar atypicalities have been reported in individuals with ASD (17), a recent study (18) conducted on a larger cohort found no difference in cerebellar anatomy between individuals with ASD and individuals with TD. Investigating cerebellar differences at the functional level might be a more promising research avenue (19). Numerous neuroimaging studies have described abnormal cerebellar activation in subjects with ASD during motor tasks and visual processing tasks (20). Despite its limited temporal resolution, the vast majority of these studies have primarily investigated cerebellar activity using functional magnetic resonance imaging (fMRI). To the best of our knowledge, EEG-based studies conducted with ASD participants focused on cortical signals and did not include the cerebellum (5). However, studies conducted in adults with TD demonstrated that both EEG and MEG can detect signals from the cerebellum (21).

In this study, we investigated the EEG of the cerebellum during an active social interaction task in ASD using the HDC paradigm. We specifically focused our analysis on the theta frequency band (3-8 Hz) (22), which has been detected in the cerebellum (23), is associated with large-scale coordination of distant brain areas (24), and might contribute to interoception, motor control, and social event anticipation, all skills critically involved in ASD (8). As previous EEG and MEG studies consistently reported atypical cortical theta power in children with ASD during resting state (25,26), we expected abnormal theta activity both at the cortical and cerebellar level in individuals with ASD.

Beyond investigating cerebellar power spectra, we also aimed at describing the connectivity patterns between the cerebellum and the sensorimotor cortex but also the prefrontal cortex (PFC) and the temporoparietal junction (TPJ). During tasks in which adults with TD had to observe repetitive hand movements, MEG analysis uncovered high connectivity between the cerebellum and the primary motor cortex in low-frequency EEG bands (< 5 Hz) (27). Additional evidence suggested that the cerebellum was connected to the prefrontal cortex (PFC) and involved in motor planning, as well as to the right TPJ (rTPJ), participating in forming inferences on others’ intentions (28). Thus, we expected atypical connectivity in the theta frequency band between the cerebellar and cerebral cortex during social interaction. To address both of our hypotheses, we used state-of-the-art source reconstruction methods to locate surface cortical and deep cerebellar sources of the recorded EEG activity.

## MATERIALS AND METHODS

### Participants

The experiment was conducted in a sample of 217 individuals: 161 had ASD and 56 had TD. Following EEG source reconstruction, we excluded 54 participants with ASD and 23 participants with TD. After preprocessing and quality check, the analyses were run on a sample of 140 participants (Table 1).

**Table 1.** Demographic and clinical characteristics of participants included in the study. Data are mean values ± standard deviation for continuous variables. ASD = Autism Spectrum Disorders; n = sample size; SRS-2 = Social Responsiveness Scale - 2nd Edition; TD = Typical Development.

|  | individuals with<br>ASD<br>(n = 107) | individuals with<br>TD<br>(n = 33) | Statistical test; p-value |
| --- | --- | --- | --- |
| Gender (male/female) | 90/16 | 18/13 | $\chi^2 = 16.11$; p = 6.71e-5 |
| Age at inclusion (years) | 13.16 ± 8.11 | 19.22 ± 9.68 | t = -3.55; p = 5.24e-4 |
| Wechsler Scale -<br>standardized total score | 102.34 ± 17.23 | 109.91 ± 13.52 | t = -2.25; p = 2.60e-2 |
| SRS-2 - Total t-score | 70.49 ± 14.16 | 45.71 ± 8.07 | t = 9.21; p = 8.14e-16 |
| Right-handedness<br>(right/left) | 85/11 | 26/4 | $\chi^2 = 0.002$; p = 0.75 |

Participants with ASD were recruited at the Child and Adolescent Psychiatry Department of the Robert Debré University Hospital, Paris (France) and were included into the study after a systematic clinical and medical examination, including negative blood test results for Fragile X syndrome. The final diagnosis of ASD was based on the Diagnostic and Statistical Manual of Mental Disorders (DSM-5) criteria and outcomes from the Autism Diagnostic Observation Schedule-Second Edition (ADOS-II) (29), the Autism Diagnostic Interview-Revised (ADI-R) (30), and the Social Responsiveness Scale–Second Edition (SRS-2) (31) for the dimensionality of social skill impairments. Participants carrying a large deletion over 2 million base pairs (Mb) detected by the Illumina 700 SNPs genotyping array and those with a medical history of epileptic seizures were excluded. The final sample for analysis only included individuals without an intellectual disability estimated with the Wechsler Intelligence Scale (32) adapted to the participant’s age. Participants with TD were recruited from the general population, and also reported no personal or familial history of ASD and/or epileptic seizures.

This study was carried out in accordance with the recommendations of the local ethics committee of the Robert Debré Hospital (study approval no. 08-029). All participants and caregivers of child participants gave written informed consent in accordance with the Declaration of Helsinki.

### Experimental Procedure

The experimental design of the study consisted of a resting state (RS) acquisition period and a social interaction task based on the HDC paradigm that all participants completed, while their neural dynamics were recorded with HD-EEG. The RS served as a control condition for comparison with the HDC experimental condition. During the RS protocol, the participant was requested to sit still and keep his/her eyes open, then closed for 30-30 seconds (conditions ‘eyes-open’ and ‘eyes-closed’). This series of two conditions was also repeated three times. The HDC protocol used a human-machine interface as initially described in Dumas et al. (9) (Figure 1). As a summary, HDC relied on a mathematical model integrating the position and speed of the participant’s index finger during movement to simulate the behavior of a VP in real-time. At the beginning of each trial, an instruction (with respect to language comprehension abilities) was given to the participant by the experimenter to synchronize index finger movement ‘in-phase’, her/his movements to those of the VP, or in ‘anti-phase’, *i*.*e*. to synchronize her/his movements with a half-period offset between the phase of the VP and the participant. Prior to the experiment, all participants were instructed that the partner was either a VP or ‘robot’ (*i*.*e*. movements will be computer-driven) or a real age- and gender-matched human peer performing the same task in another room of the hospital. The protocol consisted of 40 trials of about 20 seconds each, divided into four blocks. The instructions to the participant stayed the same within each block and alternated across blocks. The instruction for the first block was randomly assigned at the beginning of the experiment to ensure randomized order of blocks across participants.

**Figure 1.**
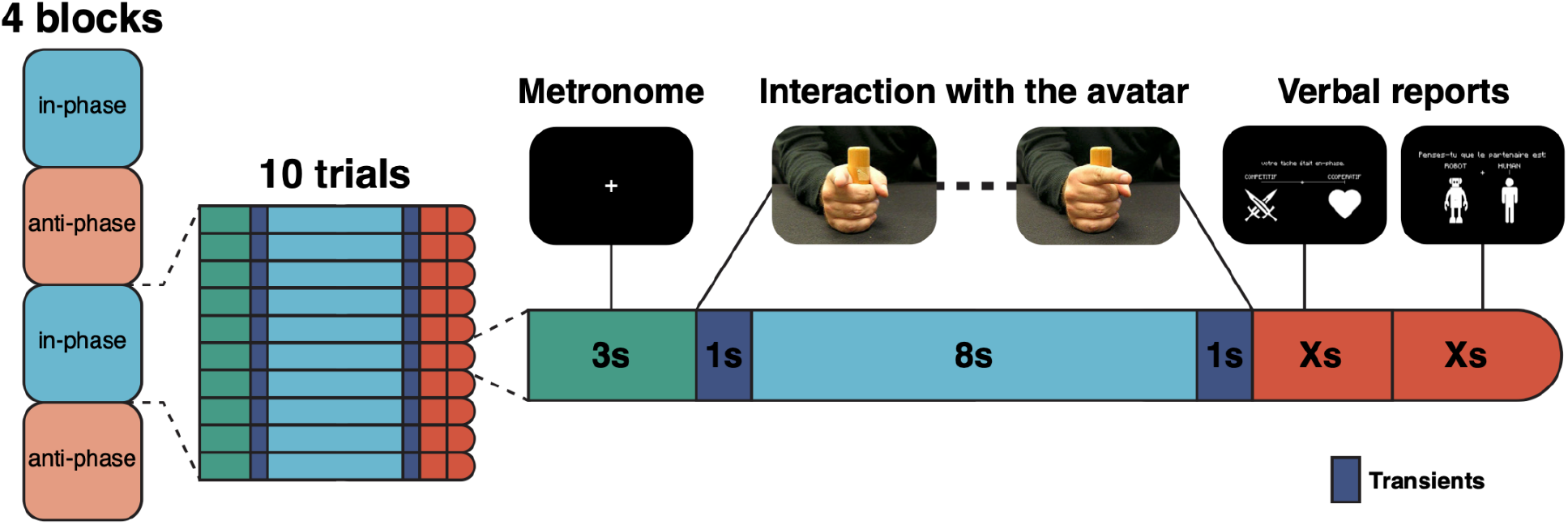
Experimental design. Structure of the protocol with four blocks alternating “in-phase/anti-phase” (order of blocks was counterbalanced across participants). Each block was divided into ten trials (left). Each trial started with a ‘warm-up’ phase in which the participant had to synchronize finger movement with the sound of a metronome for 3 seconds. Then, the participant interacted with the avatar according to the instruction (*i*.*e*. here, “in-phase”). The 1-second transient periods prevented integrating non-stationary dynamics when switching between the metronome and interaction periods. At the end of the trial, the participant provided a report for two questions displayed on the screen via finger movement: these included the impression of the participant on the competitive or cooperative behavior of the avatar and his/her impression on the human nature of the avatar. We present screenshots of what participants could see (top).

### HD-EEG acquisition

HD-EEG was recorded by a 128-channel Geodesic EEG System 400 (GES 400) with NetStation EEG software and a 128-electrode HydroCel Geodesic Sensor Net (EGI, 2017). The sensor net was connected to a Net Amps 400 amplifier (EGI, 2017), which was set to ‘255 encoding mode’ for recording external Digital Input (DIN) events (triggers). EEG data were amplified, low-pass filtered at 500 Hz, sampled at a frequency of 1000 Hz, and digitized. For the RS protocol, an event list was created in the EEG recording software with a label for each condition, which was manually selected before each task by the experimenter. For the HDC protocol, DIN events were recorded automatically. After the HD-EEG recording, the EEG electrode locations were digitized using a stereo-camera tracking digitizer (GeoScan Sensor Digitization Device, 397 Philips Neuro Inc., Eugene, OR).

### Data preprocessing

HD-EEG data were preprocessed offline with the open-source software package MNE-Python (33) to attenuate noise and artifacts from environmental and biological sources. Preprocessing started with defining events based on markers (triggers). Next, data were band-pass filtered to restrict the signal to the frequency range of interest (from delta [1-4 Hz] to gamma [30-45 Hz] waves), between 0.5 and 48 Hz. For that, a fast Fourier transform (FFT) based Finite Impulse Response (FIR) filter was used with a Hamming window, with 0.1 and 1 Hz transition bandwidths at the low and high cut-off frequencies, respectively. To avoid signal contamination from the reference or any malfunctioning channel, the signal was averaged and referenced using the Signal-Space Projection method. Then, raw data were segmented into 1-second-long epochs from both protocols (HDC, RS), epochs were decimated by a factor of 4 and the automated artifact rejection algorithm Autoreject was applied to reject or repair noisy epochs, based on channels’ trial-wise peak-to-peak amplitude (34). When the peak-to-peak signal amplitude exceeded a data-specific threshold, epochs with noisy signals were rejected. When possible, data from these epochs were interpolated and replaced within the whole length of the signal. The algorithm was first trained on epochs from both protocols (HDC, RS). Afterwards, for each protocol, relevant epochs were extracted and cleaned by the previously trained algorithm according to the above parameters.

### Sensor Analysis

To further assess the quality of the EEG data, we checked for the accuracy of our data to replicate the most robust findings from the literature. Namely, we expected the Power Spectral Density (PSD) in the alpha band (8-13 Hz) to increase in the ‘eyes-closed’ condition. We also expected to replicate a mu suppression (*i*.*e*. reduction of the oscillations in the alpha frequency range over the sensorimotor cortex in a social context) in the upper alpha band (10-13Hz) during the HDC task (35,36). In our study, we similarly observed an increased activity in the alpha band over the whole scalp, both in participants with ASD and TD; this increase of alpha power was significantly lower in the prefrontal cortex of children with ASD compared to those with TD (Sup. Figure 1). During the HDC task, in the context of social interaction, mu activity was suppressed over the scalp in both groups with TD and ASD (Sup. Figure 2). No statistically significant difference in mu activity was observed between groups.

### Source reconstruction

We reconstructed the location and spectral content of sources in the brain, using MNE-Python (33). Sources were reconstructed on an MNE-Python MRI template brain using a mixed source space. The noise-covariance matrix was then calculated from the RS ‘eyes-open’ condition with shrinkage regularization. Proper estimation of the covariance matrix is one of the most critical factors in the accuracy of the reconstruction (37). The inverse solution was computed using the Exact Low Resolution Brain Electromagnetic Tomography (eLORETA) algorithm (38).

To assess the quality of source estimation, power analysis in the alpha band during the ‘eyes-closed’ condition was replicated over the cortex after source reconstruction performed by both Minimum Norm Estimate (MNE) and eLORETA (39) algorithms (Sup. Figure 1 and Sup. Figure 3, respectively). In addition, PSD comparison in the alpha band between HDC and RS was performed over the cortex. This allowed us to check if mu suppression was also replicated at the source level, hence providing another way of validating the results of source reconstruction. This mu suppression analysis was performed on signals reconstructed with a noise covariance matrix low-pass filtered at 8 Hz in order to include only low frequencies in the noise model and avoid intrinsic alpha brain activity to be considered as noise. After source reconstruction with eLORETA, we observed that the alpha power increased significantly over the occipital lobe in both groups with ASD and TD in the ‘eyes-closed’ condition (Sup. Figure 1). No cluster was specifically associated with ASD. Similar results were observed after source reconstruction using the MNE algorithm (Sup. Figure 3). We then compared EEG signals during the social interaction (HDC task) vs resting state (RS ‘eyes-open’ condition). We observed a significant alpha power decrease all over the cerebral cortex in individuals with ASD and TD (Sup. Figure 2). We however reported no clusters specifically associated with ASD.

### Functional Connectivity

Following source reconstruction, source time courses were averaged inside regions of interest (ROIs) following the Desikan-Killiany atlas (40). Functional connectivity was calculated using the coherence metric (41) between pairs of ROIs (with 125 FFT length, and 62 overlap between segments):

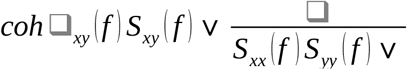

With x and y the indices of two ROIs, *f* the frequency of interest, *S*_*xy*_ the cross-spectral density between x and y, *S*_*xx*_ and *S*_*yy*_ the power spectrums of x and y (respectively).

For each subject, PSD and coherence values were averaged over the frequency band of interest and Z-scored over epochs to represent the statistical distance between conditions and account for intra-subject variability. The Z-coherence was defined as:

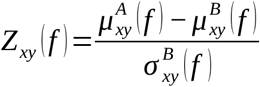

with 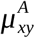 and 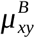 the average coherence between x and y over epochs in A (HDC) and B (RS ‘eyes-open’) conditions, and 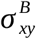 the standard deviation of the coherence in condition B over epochs.

### Statistical analysis

The power spectra of the cortex and the cerebellum were compared between groups and between conditions both at sensor and source level. Cluster-based permutation tests were used to correct for multiple comparisons over the entire brain surface (42,43). Cluster permutation tests combined spatially neighboring voxels into clusters and computed a cluster-level statistic. Permutations (n = 2000) randomly exchanged the labels of participants to estimate the distribution of the maximum cluster statistics under the null hypothesis of no difference between groups or conditions. A cluster was considered statistically significant if its statistic was greater than 95% of the maximum cluster statistics that would be expected by chance. We also used threshold-based free cluster enhancement (TFCE) correction (44), which enhanced areas of the signal belonging to cluster-like regions. Effect sizes (Z-scores for within-group tests and Cohen’s d for between-group tests) were also computed for each analysis.

Functional connectivity between the cerebellum and cortical areas during social interaction was analyzed at the source level. Functional connectivity in the theta band between the cerebellum and three cortical areas (sensorimotor cortex, PFC and TPJ) during interaction with the HDC was compared between groups using Student t-tests with false discovery rate (FDR) correction for multiple comparisons. In power and connectivity analyses, effect sizes were computed for intra- and inter-group comparisons to reduce the unequal sample size bias. Test statistics were displayed for intra-group comparisons whose results were statistically significant.

Based on the sample size of the two groups (N_ASD_ = 107; N_TD_ = 33), we calculated the statistical power of the difference between two independent means (two-tails) using the software G*Power (45). With a significance threshold of alpha = 0.05 and an estimated effect size from the literature of d = 0.5 (46), we obtained a statistical power of 1-beta = 0.7.

## RESULTS

### EEG power analysis

To ensure the robustness of the source reconstruction, Cohen’s d effect size was calculated for each dipole location and each participant between the HDC and RS conditions in the theta band. Those with more than 5% sources with a Cohen’s d value above 10 were considered as outliers. We finally ran the analysis on 140 participants, *i*.*e*. 107 with ASD and 33 with TD.

In the power analysis, we observed a decreased theta power over the right and left parietal cortex in participants with TD but not in those with ASD during social interaction (Figure 2). We counted no statistically significant clusters in the TD group. By contrast, we observed a significant increase in theta power in the temporal and frontal areas in the ASD group (TFCE value at peak = 72.68; p-value at peak = 0.01). However, we did not find any statistically significant clusters when comparing ASD and TD groups. Finally, we observed a significant increase in theta power in both cerebellar hemispheres in participants with ASD (TFCE value at peak = 33.77; p-value at peak = 5e-4) (Figure 3). We found no statistically significant clusters associated with TD.

**Figure 2.**
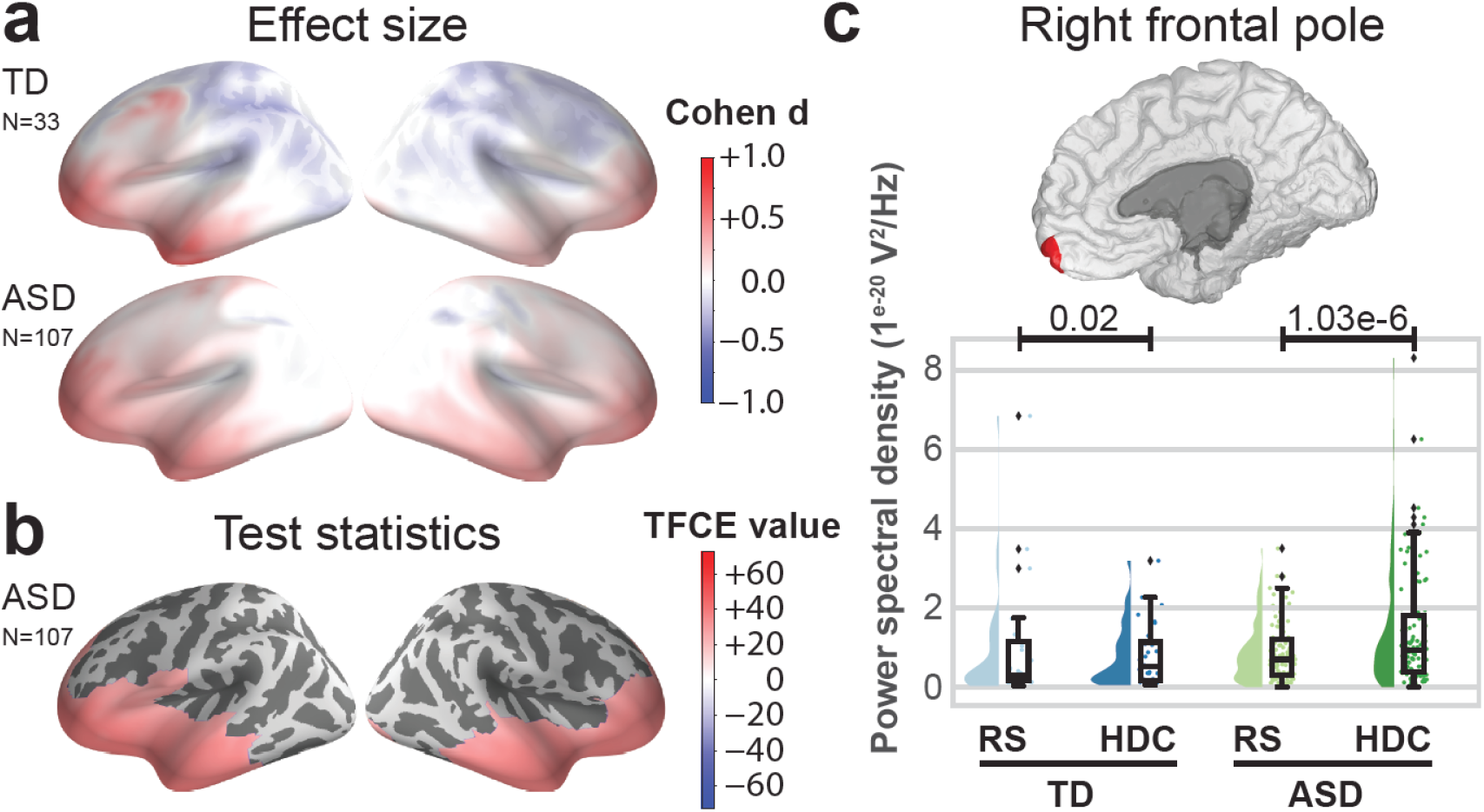
During the HDC task, cerebral cortex effect sizes and statistical maps are compared to the ones obtained in the RS ‘eyes-open’ condition for the theta band (3-8 Hz). Panels (a) and (b) show brain plots displaying the activity of the left and right hemispheres. Panel (a) shows effect sizes for groups with TD (upper) and with ASD (lower). Positive effect sizes indicate theta enhancement during the HDC. Panel (b) shows TFCE values in the group with ASD thresholded at p < 0.05 corrected. Positive values indicate greater theta enhancement during the HDC. On panel (c), half violin plots display PSD values in the right frontal pole in the TD (blue) and ASD (green) groups during RS (light) and HDC (dark) (40). For each group and condition, a density plot and a boxplot are displayed on the left and right (respectively). P-values indicate the results of intra-group comparisons (thresholded at p < 0.05, non-corrected).

**Figure 3.**
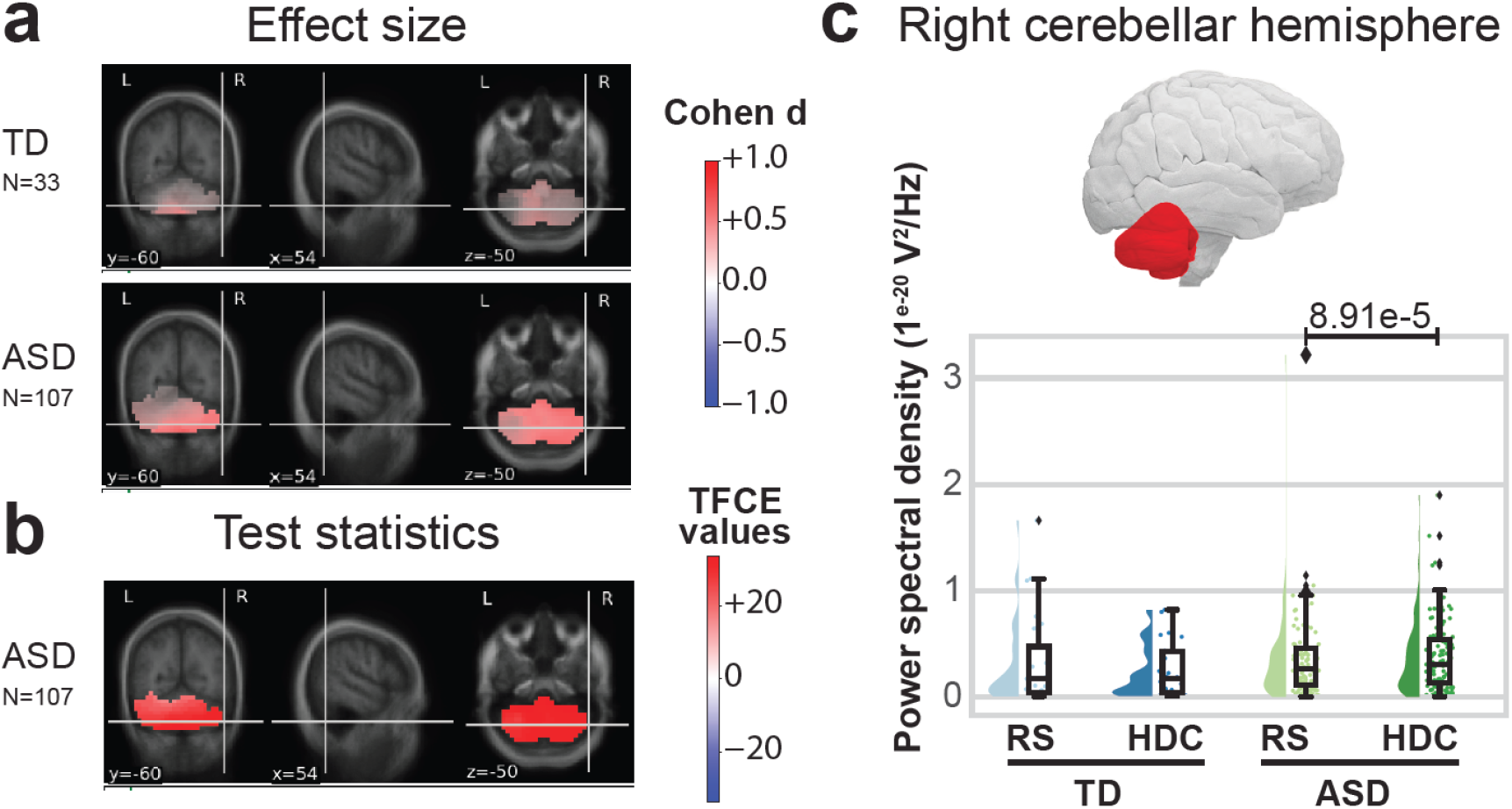
During the HDC task, the cerebellum cortex effect sizes and statistical maps are compared to those obtained in the RS ‘eyes-open’ condition for the theta band (3-8 Hz). On panel (a) and (b), the cerebellar activity is displayed on the coronal (left), sagittal (middle), and transversal (right) sections. Panel (a) shows effect sizes for the TD group (upper) and ASD group (lower). Positive effect sizes indicate theta enhancement during the HDC in the TD and ASD groups (respectively). Panel (b) shows TFCE values in the group with ASD thresholded at p < 0.05, corrected. On panel (c), half violin plots display PSD values in the right cerebellar hemisphere in the TD (blue) and ASD (green) groups during RS (light) and HDC (dark). For each group and condition, a density plot and a boxplot are displayed on the left and right (respectively). P-values indicate the results of intra-group comparisons (thresholded at p < 0.05, non-corrected).

### Connectivity analysis

We explored the connectivity between the cerebellar cortices and three main brain areas: the PFC and the TPJ, which both showed an increased power activity in the theta band during the social interaction task, and the sensorimotor cortex. We observed statistically significant interactions between the two cerebellar hemispheres and these cortical brain areas in individuals with ASD but not in those with TD (Figure 4). Compared to individuals with TD, those with ASD showed a statistical trend for an increased connectivity between the right cerebellar cortex and the ipsilateral sensorimotor cortex, and between the cerebellar cortices and the right prefrontal cortex, along with a decreased connectivity between the cerebellar cortices and the bilateral TPJs.

**Figure 4.**
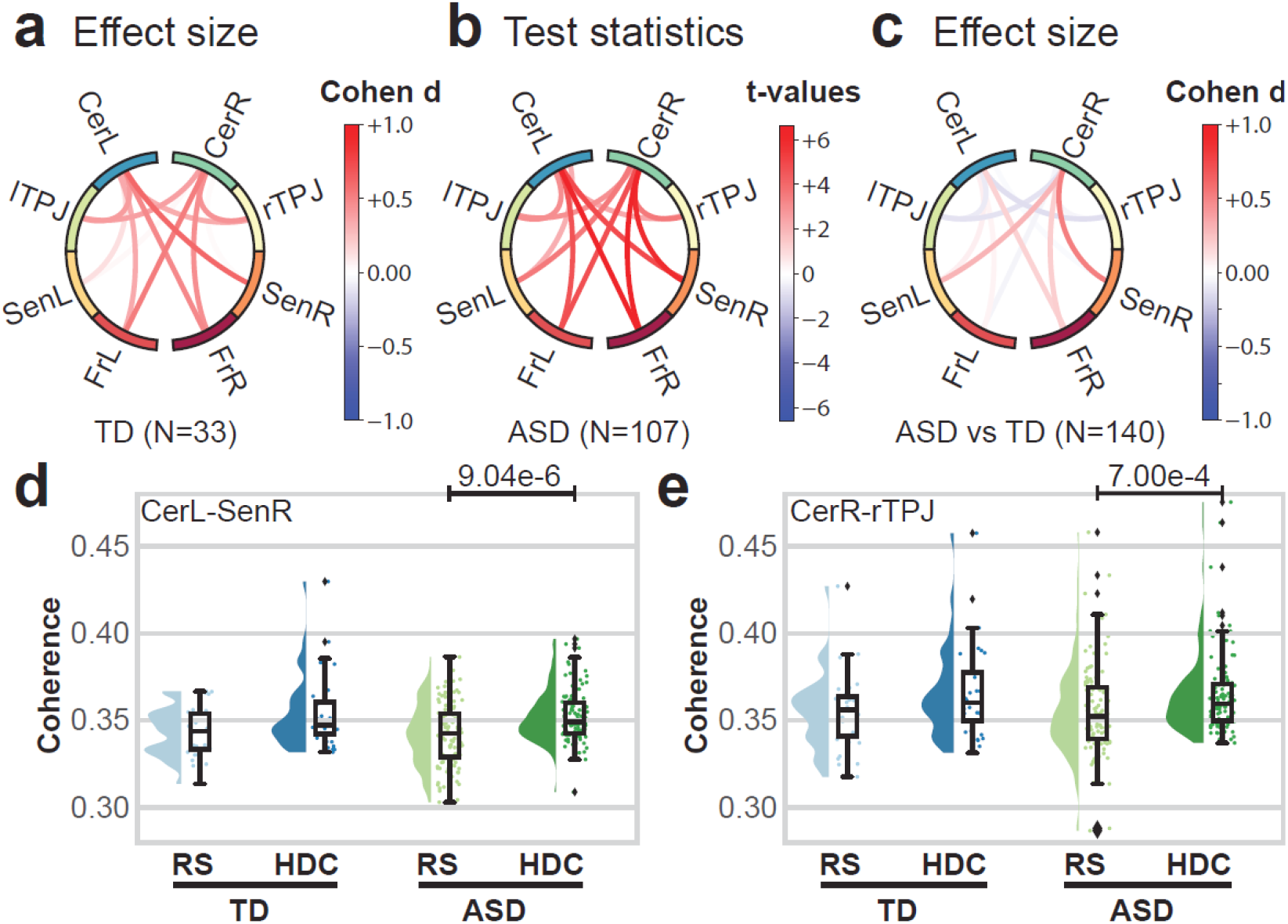
During the HDC task, the connectivity effect sizes and statistical plots were compared to those obtained in the RS ‘eyes-open’ condition for the theta band (3-8 Hz). Panels (a), (b) and (c) display a circle plot with connections between different brain areas, from top to bottom: cerebellar cortices (CerL/CerR), temporo-parietal junctions (lTPJ/rTPJ), sensorimotor cortices (SenL/SenR), and frontal cortices (FrL/FrR). Panels (a) and (c) show effect sizes for the TD and ASD *vs* TD groups (respectively). Panel (b) shows connectivity T-values for the group with ASD thresholded at p < 0.05, FDR corrected. On panel (d), half violin plots display connectivity values between the left cerebellar cortex and the right sensorimotor cortex in the TD (blue) and ASD (green) groups during RS (light) and HDC (dark). On panel (e), half violin plots display connectivity values between the right cerebellar cortex and the rTPJ in the TD (blue) and ASD (green) groups during RS (light) and HDC (dark). For each group, a density plot and a boxplot are displayed on the left and right (respectively). P-values indicate the results of intra-group comparisons (thresholded at p < 0.05, non-corrected).

## DISCUSSION

In this study, we addressed the methodological challenge of probing the role of the cerebellum in ASD during a social interaction task. By using advanced source localization, we reported an increase of theta power in the cerebellum and in the frontal and temporal cortices, associated with an increased cortico-cerebellar connectivity in individuals with ASD but not in those with neurotypical development.

The first achievement of our study was to localize cerebellar sources based on HD-EEG recordings which was considered highly challenging (21). We detected increased theta band power in the right hemisphere of the cerebellar cortex, *i*.*e*. ipsilateral to motor movement. These findings are consistent with those from preliminary studies with neurotypical participants, in which the contralateral sensorimotor cerebral cortex and the ipsilateral cerebellar cortex synchronically oscillated at the frequency of movement (47). As our study was conducted during a social interaction task, comparisons with previous studies in the literature are limited. However, our findings on significant theta power increase in the frontal and temporal cortices in ASD align with a study showing higher theta band activity in the frontal cortex of individuals with ASD vs TD during a cooperative tapping task (48). Our work further consolidates previous findings showing an excess of theta power over frontal regions and the right posterior cortex of participants with ASD during RS or non-social cognitive tasks (5). The increased cerebellar theta power we also observed in our study was only significant in individuals with ASD. This pattern, uncovered during a standardized social interaction task, is consistent with the role of the cerebellum as a regulator of social behaviors (49), and with its involvement in social cognition impairments in ASD (50). We similarly observed a significant theta power increase over frontal and temporal areas in individuals with ASD, which fosters the hypothesis of a ‘social brain network’ impairment in ASD (51). Our results stress further the diversity of brain structures involved in ‘low-level’ social interactions, such as a motor synchronization task.

The second achievement of our study was to explore the cortico-cerebellar connectivity during a social interaction task using the HDC paradigm. We observed an increased connectivity between the cerebellum and all cortical brain ROIs in both groups, which reached significance only in participants with ASD. Our results reinforce previous findings in adults with neurotypical development undergoing the same HDC protocol (9). Based on effect size comparisons, we reported that cortico-cerebellar connectivity (specifically between the cerebellum and the sensorimotor and frontal cortices) in participants with ASD was higher during social interaction than in those with TD. Interestingly, the hyperconnectivity in the theta frequency between the ipsilateral sensorimotor cortex and the right cerebellar cortex we reported in ASD displayed an opposite trend in controls. Again, our results are in line with previous resting state fMRI studies, which showed hyperconnectivity patterns between the cerebellum and sensorimotor areas in participants with ASD compared to controls (3,52). Effect size comparisons in our study revealed a lower connectivity between the cerebellum and the rTPJ in participants with ASD than in controls during social interaction. These results are supported by several resting state fMRI studies, which report weak connectivity patterns between the cerebellum and supramodal cortical regions, including the rTPJ in ASD during resting state and social mentalizing tasks (3,53). The hypoconnectivity between the cerebellum and the rTPJ could represent a relevant biomarker of social interaction impairment, related to sensorimotor integration dysfunction (11).

Our study should be considered in light of its limitations. First, the low number of participants with TD might account for the non-significant results we reported when performing within-group and group-level comparisons (statistical power = 0.7 for an effect size of 0.5). Second, participants with ASD were on average younger than controls, and the sex ratio was unbalanced in the ASD group, which included more male than female participants. Considering that altered connectivity findings in ASD are not uniform across age and sex (54), some of the atypical neural dynamics we reported might arise from a sampling bias in our study. Third, we used a normalized adult brain template to estimate the source localization. This might have created a bias as we did not consider the variability linked to the size of the brain across ages but also the neuroanatomical heterogeneity associated with ASD (55). Similarly, we excluded from the analysis a large number of participants (n = 77) by using a stringent algorithm to detect EEG artifacts. This may also create a sampling bias in our study since it excludes subjects with significant comorbid hyperactivity. The development of new tools for artifact correction would enable us to include an increased number of data segments, limit the sampling bias and increase the statistical power of the study. In conclusion, our results provided additional support for tracking the cortico-cerebellar dynamics during real-time social interaction in a clinical pediatric setting. The study uncovered new functional brain patterns associated with ASD and stressed further the role of theta oscillations in social interaction. Our study paves the way for multimodal interactive psychometrics for the fine-grained assessment of social skills and their underlying neural mechanisms in ASD during naturalistic dynamic interactions (12,56).

## Data Availability

All data produced in the present study are available upon reasonable request to the authors.

## ACKNOWLEDGMENTS

The authors would like to thank all participants and their families who participated in the study, and the Clinical Investigation Center, Robert Debré Hospital, Paris, France for the management of the study. They also thank Karim Jerbi and Julie Grézes for their constructive comments on an early version of the manuscript. This study was supported by funding from Fondation de France (2015-00059547) and Institut Pasteur. A.B. was supported by a fellowship of the French Ministry of Foreign Affairs. G.D. was supported by the Institute for Data Valorization (IVADO) Professor Startup & Operational Funds and by Fonds de la Recherche en Santé du Québec (FRSQ) junior 1 salary award.

## SUPPLEMENTARY FIGURE LEGENDS

**Sup. Figure 1.**
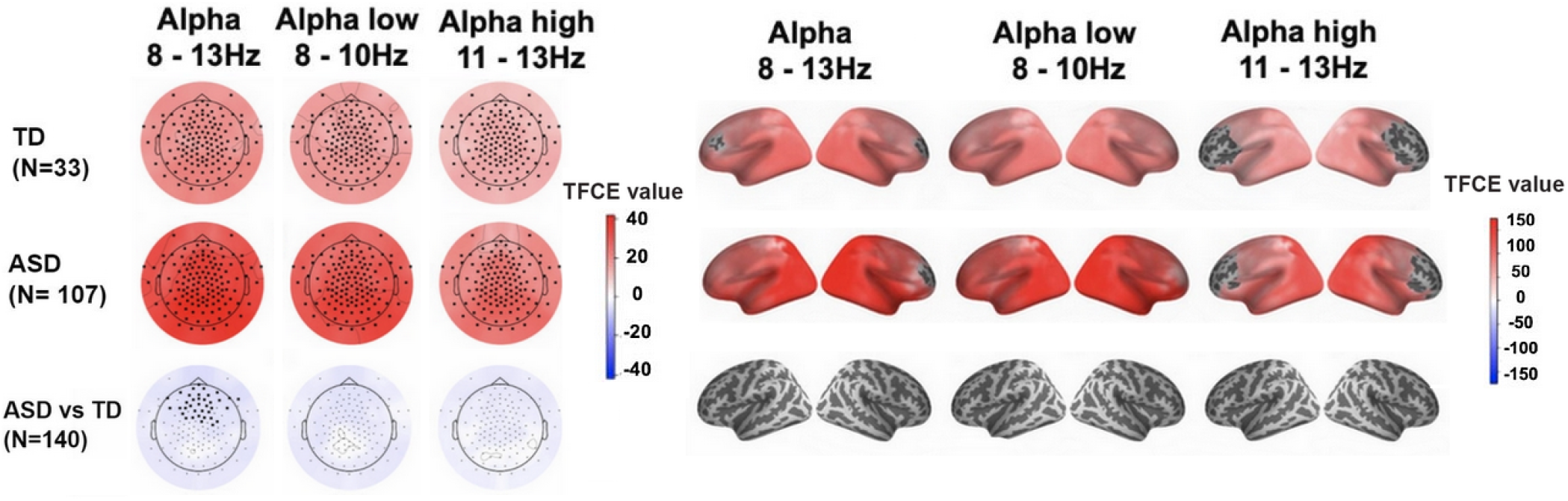
Scalp (left panel) and cerebral cortex (right panel) statistical maps during RS ‘eyes-closed’ compared to the RS ‘eyes-open’ condition for the alpha band (8-13 Hz), low alpha band (8-10 Hz) and high alpha band (10-13 Hz). In both panels, the upper, middle and lower rows show TFCE values for the TD and ASD groups, and the ASD *vs* TD groups comparison (respectively), thresholded at p < 0.05, TFCE corrected. In the upper and middle rows, positive TFCE values indicate alpha enhancement during RS ‘eyes-closed’ vs RS ‘eyes-open’ in the TD and ASD groups (respectively). In the lower row, negative TFCE values indicate lower alpha enhancement during RS ‘eyes-closed’ in the ASD *vs* TD groups.

**Sup. Figure 2.**
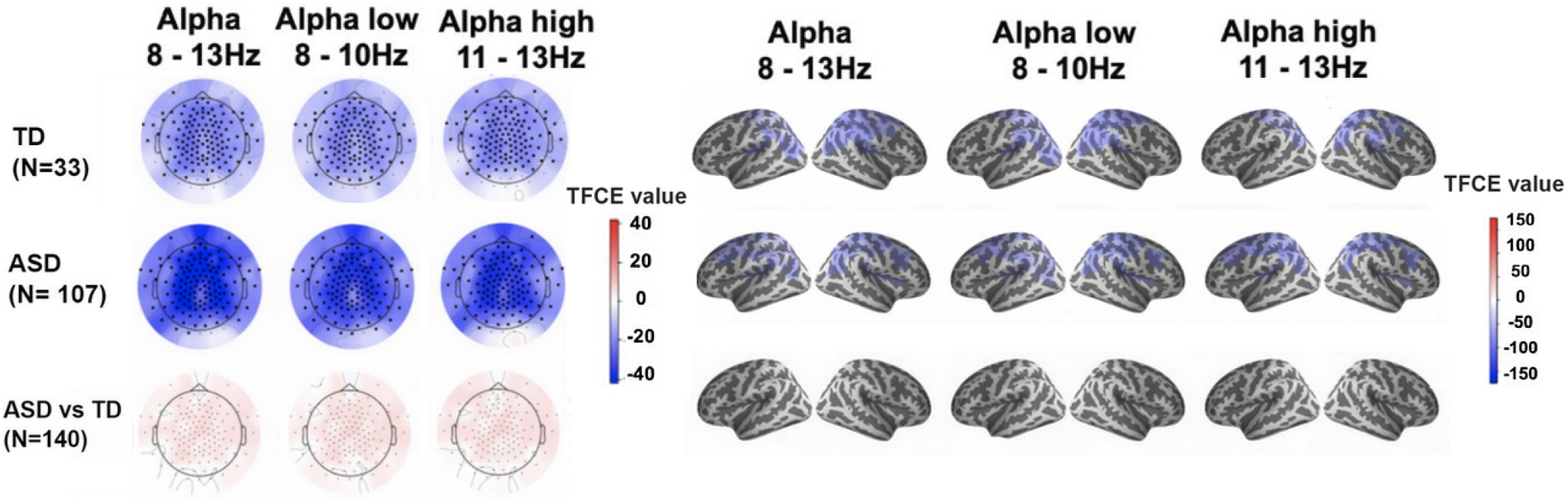
Scalp (left panel) and cerebral cortex (right panel) statistical maps during the HDC compared to the RS ‘eyes-open’ condition for the alpha (8-13 Hz), low alpha (8-10 Hz) and high alpha (11-13 Hz) bands. In both panels, the upper, middle and lower rows show TFCE values for the TD and ASD groups, and the ASD *vs* TD groups comparison (respectively), thresholded at p < 0.05, TFCE corrected. In the upper and middle rows, negative TFCE values indicate alpha suppression during HDC *vs* RS in the TD and ASD groups (respectively). In the lower row, positive TFCE values indicate greater alpha enhancement during the HDC task in the ASD *vs* TD groups.

**Sup. Figure 3.**
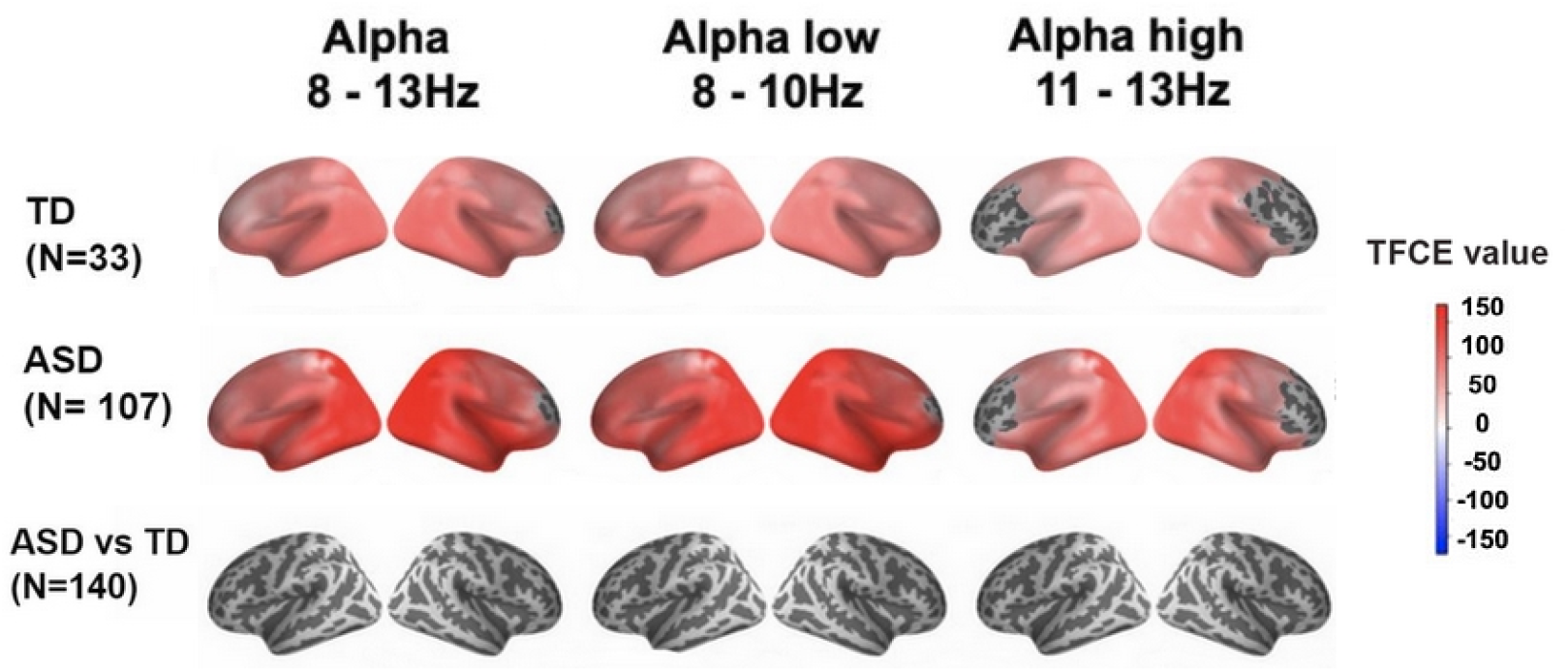
Cerebral cortex statistical maps during RS ‘eyes-closed’ compared to the RS ‘eyes-open’ condition for the alpha band (8-13 Hz), low alpha band (8-10 Hz) and high alpha band (11-13 Hz) after source reconstruction with the MNE algorithm. Statistical maps are similar to that obtained after source reconstruction with the eLORETA algorithm (Sup. Figure 1, right panel). The upper, middle and lower rows show TFCE values for the TD and ASD groups, and the ASD *vs* TD group comparison (respectively), thresholded at p < 0.05, TFCE corrected. In the upper and middle rows, positive TFCE values indicate alpha enhancement during RS ‘eyes-closed’ vs RS ‘eyes-open’ in the TD and ASD groups (respectively). In the lower row, negative TFCE values indicate lower alpha enhancement during RS ‘eyes-closed’ in the ASD *vs* TD groups.

